# Genomic analysis of early spread of monkeypox virus in Washington State

**DOI:** 10.1101/2022.09.19.22280115

**Authors:** Pavitra Roychoudhury, Jaydee Sereewit, Hong Xie, Ethan Nunley, Nicole A.P. Lieberman, Alexander L. Greninger

**Author notes:** Corresponding author: Pavitra Roychoudhury, 1100 Fairview Ave N, E5-110, Seattle WA 98109-1024, /. contributed equally (Pavitra Roychoudhury, Jaydee Sereewit).

## Abstract

Genomic analysis of the monkeypox virus outbreak in Washington State using 109 viral genome sequences collected from July-August 2022 shows low overall genetic diversity, multiple introductions into the state with ongoing community transmission, and potential for coinfection of an individual by multiple strains.

**Biography:** Dr. Roychoudhury is Acting Instructor and Director of COVID-19 NGS in the Department of Laboratory Medicine and Pathology at the University of Washington and Associate in the Vaccine and Infectious Disease Division at the Fred Hutchinson Cancer Research Center. Her primary research interests are pathogen genomics and mathematical models of viral evolution and host-pathogen interactions.

## Main text

The World Health Organization declared the monkeypox outbreak of 2022 a public health emergency of international concern on 23-Jul-2022 after cases were identified in nearly 80 countries (1). As of 26-Aug-2022, there were 411 confirmed cases of monkeypox in Washington state (https://doh.wa.gov/you-and-your-family/illness-and-disease-z/monkeypox, accessed 28-Aug-22), and 17,432 cases in the United States (https://www.cdc.gov/poxvirus/monkeypox/response/2022/us-map.html, accessed 28-Aug-22). Viral whole genome sequencing (WGS) is a valuable tool in the outbreak response toolkit because it can augment contact tracing efforts and identify the emergence of new variants with potential impacts on infectivity, virulence, vaccine escape and treatment resistance. To date, Washington state has deposited the largest number of monkeypox sequences in the United States (Supp table 1). Here we describe the Washington outbreak using 109 monkeypox virus (MPXV) genomes from the state.

WGS was attempted on residual clinical specimens (primarily lesion swabs) that were positive for MPXV by PCR with a cycle threshold (Ct) value under 31 (range 15.9 to 30.4, n = 140). Sequencing was performed using a hybridization probe-capture based approach as previously described, with probes designed based on the MXPV 2022/MA001 strain sequence (Supplementary Methods) (2). Consensus genomes were generated using a custom pipeline that performs trimming, filtering, and iterative remapping (https://github.com/greninger-lab/revica, Supplemental Methods). Sequences with < 1% Ns were deposited to GenBank under PRJNA862948 (accessions in Supplementary Table 1). Phylogenetic analysis was performed using Augur, Auspice, and Nextclade (3,4). This study was approved by the University of Washington Institutional Review Board.

A total of 109 sequences were included in this analysis, obtained from 98 individuals with specimen collection dates between 6-Jul-2022 and 19-Aug-2022, primarily from King and Pierce counties. Of the 98 individuals, 90 were male (86.0%), 1 female (1.1%), and 7 unknown/undeclared sex (11.8%). Median age at specimen collection was 36.0 years (range 19 to 57 years).

We identified multiple identical genomes from different individuals, suggesting ongoing community transmission (Figure 1A). All 109 genomes fell within the predominant 2022 outbreak lineage B.1 (5), and included sub-lineages B.1.1 (n = 18), B.1.2 (n = 6), B.1.3 (n = 10), B.1.4 (n = 2), and B.1.8 (n = 2) suggesting separate introductions into the state. Overall, we observed low genetic diversity with a median of 1 amino acid mutations (substitutions or deletions) across the genome (range 0 to 7) relative to the B.1 ancestor (reference MPXV_USA_2022_MA001/ Genbank ON563414). There were 138 unique single nucleotide polymorphisms (SNPs) identified across the genome in the 109 sequences (Figure 1B), producing 66 unique mutations (amino acid substitutions or deletions) in 51 genes. Five unique amino acid substitutions (A1232V, D1546N, D1604N, S1633L, S553N) occurred in surface glycoprotein OPG210, and three (D441Y, E306K, E553K) in OPG189 which encodes one of several ankyrin-repeat proteins, and three (S553N, A1232V, D1604N) in surface glycoprotein OPG210 (Figure 1C). There was an abundance of G to A and C to T nucleotide substitutions (Figure 1D), indicative of Apolipoprotein B mRNA Editing Catalytic Polypeptide-like3 (APOBEC3) activity and consistent with other reports (6). No substitutions or deletions were identified in OPG057, a membrane glycoprotein homologous to F13L in Vaccinia virus and the putative target of the therapeutic antiviral tecovirimat currently in use (7).

**Figure 1.**
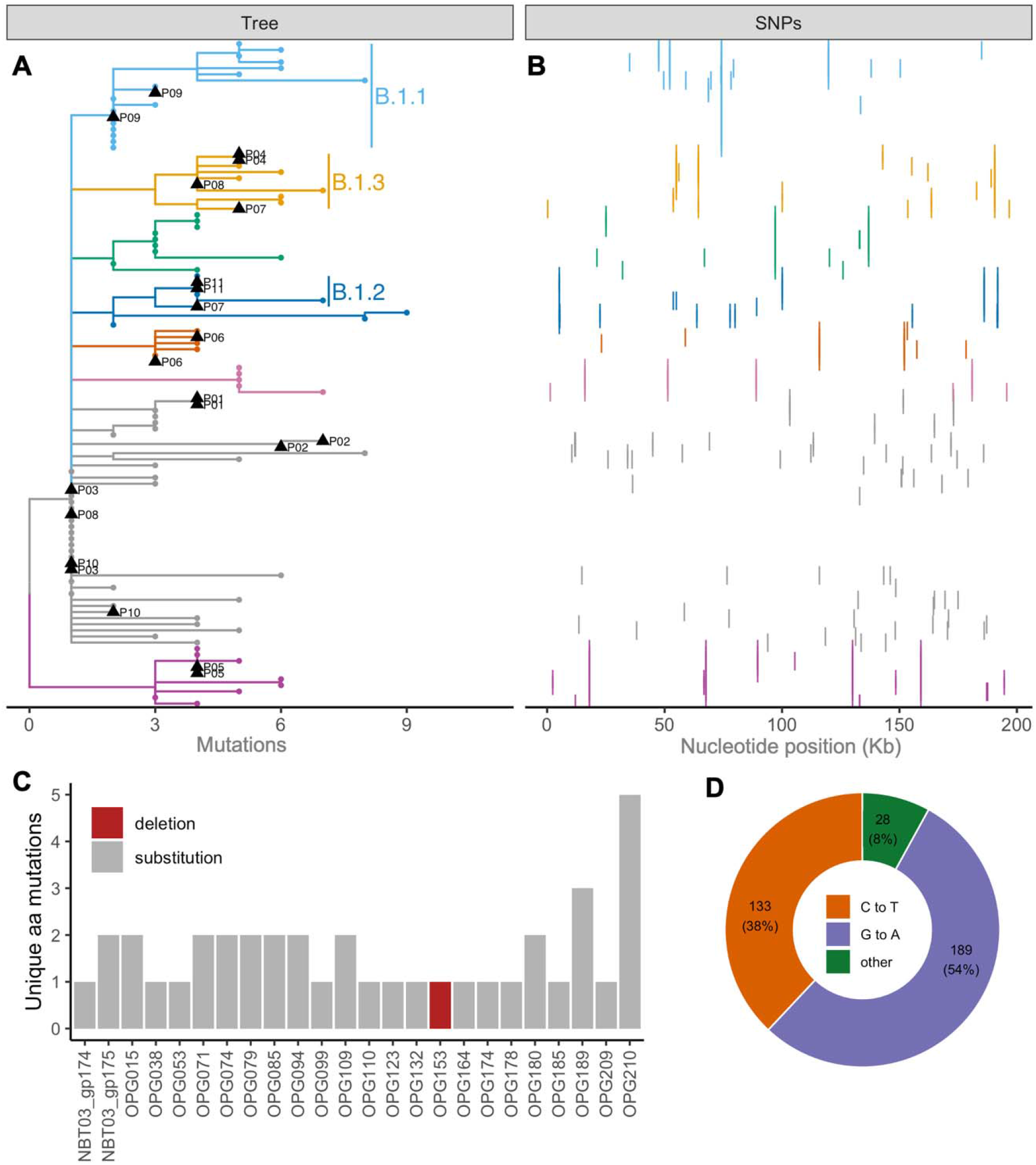
**A)** Phylogenetic analysis of 109 MPXV genomes shows that all Washington sequences fall within the major outbreak lineage B.1, with many identical sequences suggesting community transmission and distinct sub-lineages suggesting multiple introductions into the state. Sequences from multiple swabs from the same individual were available for 11 individuals (P01 to P11, tips indicated with black triangles). **B)** Single nucleotide polymorphisms from each sample in panel A arrayed across the MPXV genome. **C)** After excluding genes with mutations (amino acid substitutions or deletions) in a single sample, most genes contained 1-2 mutations, with the exception of OPG189 and OPG210. **D)** There was an abundance of G to A and C to T mutations, indicating likely APOBEC3 involvement as reported by others (6).

Sequences from multiple swabs from the same person at the same time point had a median pairwise nucleotide difference of 1 (range = 0 to 10, n = 11 sample pairs) outside of labile tandem repeat regions (8). Even greater similarity was observed in protein sequences with 0 median pairwise amino acid differences (range = 0 to 6 differences). Three sample pairs (from individuals P06, P07 and P08) had 1, 6 and 2 amino acid differences respectively. Relative to the B.1 ancestral strain, one of the P06 pair featured a V195I mutation in OPG079. One of the P08 pair had synonymous mutations in OPG073 and OPG083, as well as an OPG003:R84K substitution. Finally, differences in repeat samples from P07 suggest possible coinfection with strains from the B.1.2 and B.1.3 lineage, consistent with the patient’s clinical history. Relative to the MA001 B.1 reference, one of the P07 samples had synonymous mutations in OPG083 and OPG189, OPG180:D325N, and OPG016:R84K. The other of the P07 pair shared none of those SNPs, but had OPG015:V261A, OPG109:I66V and the B.1.2-defining OPG210:D1604N. These mutations remained after re-extracting and re-sequencing the original specimens and, when compared to interhost variation, suggest the possibility of co-infection with different MPXV strains.

Overall, our data show ongoing community transmission of monkeypox in Washington. While the limited genetic diversity makes it challenging to use sequence data for contact tracing, continued genomic surveillance will be crucial for tracking viral evolution and early identification of mutations associated with vaccine escape or antiviral treatment resistance.

## Data Availability

All sequencing data in this manuscript has been deposited to public databases NCBI Genbank and SRA with accessions included in the manuscript, code is available on Github at https://github.com/greninger-lab/revica.

## Supplementary Methods

### Sequencing approach

DNA was extracted using the MagNA Pure 96 DNA and Viral NA Small Volume Kit (Roche), and sequencing libraries were prepared using the DNAPrep kit (Illumina) and a custom xGEN NGS Hybridization Capture DNA panel (IDT) based on MXPV 2022/MA001 strain sequence (NCBI GenBank accession ON563414.2). Libraries were sequenced on Illumina Nextseq 2000 or NovaSeq 6000 instruments using 2×150bp kits targeting at least 1 million reads per sample.

### Bioinformatic Analysis

Paired-end raw reads were adapter- and quality-trimmed with Trimmomatic (v0.39). Unpaired reads and reads shorter than 120bp were discarded. Trimmed reads were aligned to the West African MPXV reference strain NC_063383.1 using bbmap (v38.96), and duplicated reads were discarded.

Ambiguously mapped reads were randomly assigned to one of the top-scoring sites to give the inverted terminal repeats regions even coverage. The consensus genome was generated by three iterations of consensus calling using Samtools mpileup (v1.15) and iVar consensus (v1.3.1) with a minimum base quality of 15, a minimum frequency threshold of 0.6, and a minimum depth of 5, followed by remapping reads to the most recent consensus. After each iteration, any leading or trailing Ns were removed. (https://github.com/greninger-lab/revica)

**Supplementary Table 1:** Sequencing data is available on NCBI Genbank and raw reads have been deposited to the NCBI sequence read archive (SRA). Patient numbers are indicated for sequences obtained from the same individual and correspond to tip labels on the phylogenetic tree in Figure 1A. Resequencing was performed to confirm coinfection on two samples and these reads are deposited in separate SRRs indicated by an asterisk.

| Isolate Name | GenBank Accession | SRA Accession | Patient Number |
| --- | --- | --- | --- |
| MpxV/human/USA/WA-UW-083698/2022 | OP442945.1 | SRR21524973 | P01 |
| MpxV/human/USA/WA-UW-088793/2022 | OP442947.1 | SRR21524979 | P01 |
| MpxV/human/USA/WA-UW-083781/2022 | OP392544.1 | SRR21524974 | P02 |
| MpxV/human/USA/WA-UW-085171/2022 | OP392546.1 | SRR21524961 | P02 |
| MpxV/human/USA/WA-UW-087006/2022 | OP257260.1 | SRR21236084 | P03 |
| MpxV/human/USA/WA-UW-088092/2022 | OP392551.1 | SRR21524968 | P03 |
| MpxV/human/USA/WA-UW-086040/2022 | OP257258.1 | SRR21236086 | P04 |
| MpxV/human/USA/WA-UW-082770/2022 | OP392540.1 | SRR21524969 | P04 |
| MpxV/human/USA/WA-UW-083953/2022 | OP328307.1 | SRR21236131 | P05 |
| MpxV/human/USA/WA-UW-087336/2022 | OP392550.1 | SRR21524987 | P05 |
| MpxV/human/USA/WA-UW-087301/2022 | OP310047.1 | SRR21236139 | P06 |
| MpxV/human/USA/WA-UW-084148/2022 | OP392545.1 | SRR21524986 | P06 |
| MpxV/human/USA/WA-UW-074949/2022 | OP184762.1 | SRR20973038<br>SRR21616210* | P07 |
| MpxV/human/USA/WA-UW-074988/2022 | OP184765.1 | SRR20973035<br>SRR21616209* | P07 |
| MpxV/human/USA/WA-UW-073669/2022 | OP123049.1 | SRR20736989 | P08 |
| MpxV/human/USA/WA-UW-076854/2022 | OP123050.1 | SRR20736988 | P08 |
| MpxV/human/USA/WA-UW-076773/2022 | OP184763.1 | SRR20973037 | P09 |
| MpxV/human/USA/WA-UW-074932/2022 | OP184764.1 | SRR20973036 | P09 |
| MpxV/human/USA/WA-UW-071121/2022 | OP123047.1 | SRR20736991 | P10 |
| MpxV/human/USA/WA-UW-076861/2022 | OP123048.1 | SRR20736990 | P10 |
| MpxV/human/USA/WA-UW-087424/2022 | OP257248.1 | SRR21236101 | P11 |
| MpxV/human/USA/WA-UW-085462/2022 | OP392547.1 | SRR21524972 | P11 |
| MpxV/human/USA/WA-UW-073909/2022 | OP055800.1 | SRR20653196 |  |
| MpxV/human/USA/WA-UW-076724/2022 | OP055804.1 | SRR20653192 |  |
| MpxV/human/USA/WA-UW-079141/2022 | OP055806.1 | SRR20653190 |  |
| MpxV/human/USA/WA-UW-074372/2022 | OP055807.1 | SRR20653189 |  |
| MpxV/human/USA/WA-UW-075986/2022 | OP055808.1 | SRR20653188 |  |
| MpxV/human/USA/WA-UW-076082/2022 | OP055809.1 | SRR20653187 |  |
| MpxV/human/USA/WA-UW-073974/2022 | OP123040.1 | SRR20731575 |  |
| MpxV/human/USA/WA-UW-070134/2022 | OP123041.1 | SRR20731574 |  |
| MpxV/human/USA/WA-UW-077622/2022 | OP123042.1 | SRR20731573 |  |
| MpxV/human/USA/WA-UW-077836/2022 | OP123043.1 | SRR20731572 |  |
| MpxV/human/USA/WA-UW-071246/2022 | OP123044.1 | SRR20736996 |  |
| MpxV/human/USA/WA-UW-076225/2022 | OP123045.1 | SRR20736995 |  |
| MpxV/human/USA/WA-UW-078870/2022 | OP123046.1 | SRR20736992 |  |
| MpxV/human/USA/WA-UW-070499/2022 | OP169336.1 | SRR20913445 |  |
| MpxV/human/USA/WA-UW-074224/2022 | OP169340.1 | SRR20913438 |  |
| MpxV/human/USA/WA-UW-074184/2022 | OP169341.1 | SRR20913437 |  |
| MpxV/human/USA/WA-UW-071966/2022 | OP169344.1 | SRR20913433 |  |
| MpxV/human/USA/WA-UW-075687/2022 | OP169345.1 | SRR20913443 |  |
| MpxV/human/USA/WA-UW-072839/2022 | OP169346.1 | SRR20913442 |  |
| MpxV/human/USA/WA-UW-079401/2022 | OP184760.1 | SRR20973040 |  |
| MpxV/human/USA/WA-UW-078603/2022 | OP184761.1 | SRR20973039 |  |
| MpxV/human/USA/WA-UW-089015/2022 | OP257243.1 | SRR21236108 |  |
| MpxV/human/USA/WA-UW-080268/2022 | OP257244.1 | SRR21236106 |  |
| MpxV/human/USA/WA-UW-084909/2022 | OP257245.1 | SRR21236104 |  |
| MpxV/human/USA/WA-UW-083578/2022 | OP257246.1 | SRR21236103 |  |
| MpxV/human/USA/WA-UW-088432/2022 | OP257247.1 | SRR21236102 |  |
| MpxV/human/USA/WA-UW-081786/2022 | OP257249.1 | SRR21236100 |  |
| MpxV/human/USA/WA-UW-087527/2022 | OP257250.1 | SRR21236099 |  |
| MpxV/human/USA/WA-UW-081272/2022 | OP257251.1 | SRR21236096 |  |
| MpxV/human/USA/WA-UW-085937/2022 | OP257252.1 | SRR21236095 |  |
| MpxV/human/USA/WA-UW-088081/2022 | OP257253.1 | SRR21236094 |  |
| MpxV/human/USA/WA-UW-080706/2022 | OP257257.1 | SRR21236088 |  |
| MpxV/human/USA/WA-UW-085684/2022 | OP257259.1 | SRR21236085 |  |
| MpxV/human/USA/WA-UW-084325/2022 | OP257261.1 | SRR21236083 |  |
| MpxV/human/USA/WA-UW-087619/2022 | OP257262.1 | SRR21236082 |  |
| MpxV/human/USA/WA-UW-089987/2022 | OP257263.1 | SRR21236081 |  |
| MpxV/human/USA/WA-UW-080976/2022 | OP257264.1 | SRR21236080 |  |
| MpxV/human/USA/WA-UW-089183/2022 | OP257265.1 | SRR21236078 |  |
| MpxV/human/USA/WA-UW-083179/2022 | OP257266.1 | SRR21236077 |  |
| MpxV/human/USA/WA-UW-084449/2022 | OP257267.1 | SRR21236142 |  |
| MpxV/human/USA/WA-UW-080617/2022 | OP310039.1 | SRR21236109 |  |
| MpxV/human/USA/WA-UW-088656/2022 | OP310040.1 | SRR21236098 |  |
| MpxV/human/USA/WA-UW-080185/2022 | OP310041.1 | SRR21236087 |  |
| MpxV/human/USA/WA-UW-089266/2022 | OP310042.1 | SRR21236076 |  |
| MpxV/human/USA/WA-UW-086841/2022 | OP310043.1 | SRR21236075 |  |
| MpxV/human/USA/WA-UW-083888/2022 | OP310044.1 | SRR21236074 |  |
| MpxV/human/USA/WA-UW-083914/2022 | OP310046.1 | SRR21236140 |  |
| MpxV/human/USA/WA-UW-088871/2022 | OP310048.1 | SRR21236138 |  |
| MpxV/human/USA/WA-UW-082029/2022 | OP310051.1 | SRR21236135 |  |
| MpxV/human/USA/WA-UW-081935/2022 | OP310052.1 | SRR21236134 |  |
| MpxV/human/USA/WA-UW-082670/2022 | OP310053.1 | SRR21236133 |  |
| MpxV/human/USA/WA-UW-088213/2022 | OP310054.1 | SRR21236132 |  |
| MpxV/human/USA/WA-UW-086360/2022 | OP310055.1 | SRR21236130 |  |
| MpxV/human/USA/WA-UW-089437/2022 | OP310056.1 | SRR21236129 |  |
| MpxV/human/USA/WA-UW-089714/2022 | OP310057.1 | SRR21236128 |  |
| MpxV/human/USA/WA-UW-087349/2022 | OP310058.1 | SRR21236127 |  |
| MpxV/human/USA/WA-UW-088764/2022 | OP310061.1 | SRR21236124 |  |
| MpxV/human/USA/WA-UW-089836/2022 | OP310062.1 | SRR21236123 |  |
| MpxV/human/USA/WA-UW-082810/2022 | OP310063.1 | SRR21236122 |  |
| MpxV/human/USA/WA-UW-082497/2022 | OP310064.1 | SRR21236121 |  |
| MpxV/human/USA/WA-UW-088973/2022 | OP310065.1 | SRR21236119 |  |
| MpxV/human/USA/WA-UW-089152/2022 | OP310066.1 | SRR21236118 |  |
| MpxV/human/USA/WA-UW-082625/2022 | OP310067.1 | SRR21236116 |  |
| MpxV/human/USA/WA-UW-088225/2022 | OP310068.1 | SRR21236105 |  |
| MpxV/human/USA/WA-UW-088152/2022 | OP310069.1 | SRR21236079 |  |
| MpxV/human/USA/WA-UW-083083/2022 | OP328308.1 | SRR21236115 |  |
| MpxV/human/USA/WA-UW-089142/2022 | OP328309.1 | SRR21236114 |  |
| MpxV/human/USA/WA-UW-088725/2022 | OP328310.1 | SRR21236112 |  |
| MpxV/human/USA/WA-UW-088258/2022 | OP328311.1 | SRR21236111 |  |
| MpxV/human/USA/WA-UW-087564/2022 | OP328312.1 | SRR21236110 |  |
| MpxV/human/USA/WA-UW-080100/2022 | OP392535.1 | SRR21524985 |  |
| MpxV/human/USA/WA-UW-080292/2022 | OP392536.1 | SRR21524966 |  |
| MpxV/human/USA/WA-UW-080834/2022 | OP392537.1 | SRR21524964 |  |
| MpxV/human/USA/WA-UW-081469/2022 | OP442944.1 | SRR21524980 |  |
| MpxV/human/USA/WA-UW-081714/2022 | OP392538.1 | SRR21524978 |  |
| MpxV/human/USA/WA-UW-082215/2022 | OP392539.1 | SRR21524967 |  |
| MpxV/human/USA/WA-UW-082488/2022 | OP442941.1 | SRR21236117 |  |
| MpxV/human/USA/WA-UW-082880/2022 | OP392541.1 | SRR21524977 |  |
| MpxV/human/USA/WA-UW-083506/2022 | OP392542.1 | SRR21524981 |  |
| MpxV/human/USA/WA-UW-083584/2022 | OP392543.1 | SRR21524963 |  |
| MpxV/human/USA/WA-UW-084331/2022 | OP442942.1 | SRR21236107 |  |
| MpxV/human/USA/WA-UW-085393/2022 | OP442946.1 | SRR21524975 |  |
| MpxV/human/USA/WA-UW-086026/2022 | OP442943.1 | SRR21236097 |  |
| MpxV/human/USA/WA-UW-087094/2022 | OP392548.1 | SRR21524960 |  |
| MpxV/human/USA/WA-UW-087104/2022 | OP392549.1 | SRR21524976 |  |
| MpxV/human/USA/WA-UW-088325/2022 | OP392552.1 | SRR21524971 |  |
| MpxV/human/USA/WA-UW-088960/2022 | OP392553.1 | SRR21524982 |  |

## References

1. Kozlov M. Monkeypox declared a global emergency: will it help contain the outbreak? Nature [Internet]. 2022 Jul 25 [cited 2022 Aug 28]; Available from: https://www.nature.com/articles/d41586-022-02054-7

2. Greninger AL, Roychoudhury P, Xie H, Casto A, Cent A, Pepper G, et al. Ultrasensitive Capture of Human Herpes Simplex Virus Genomes Directly from Clinical Samples Reveals Extraordinarily Limited Evolution in Cell Culture. mSphere. 2018 Jun 13;3(3):1–12.

3. Aksamentov I, Roemer C, Hodcroft E, Neher R. Nextclade: clade assignment, mutation calling and quality control for viral genomes. J Open Source Softw. 2021 Nov 30;6(67):3773.

4. Hadfield J, Megill C, Bell SM, Huddleston J, Potter B, Callender C, et al. Nextstrain: real-time tracking of pathogen evolution. bioRxiv. nov; 2017.

5. Wang L, Shang J, Weng S, Aliyari SR, Ji C, Cheng G, et al. Genomic annotation and molecular evolution of monkeypox virus outbreak in 2022. J Med Virol [Internet]. 2022 Jul 29; Available from: http://dx.doi.org/10.1002/jmv.28036

6. Gigante CM, Korber B, Seabolt MH, Wilkins K, Davidson W, Rao AK, et al. Multiple lineages of Monkeypox virus detected in the United States, 2021-2022 [Internet]. bioRxiv. 2022 [cited 2022 Sep 9]. p. 2022.06.10.495526. Available from: https://www.biorxiv.org/content/10.1101/2022.06.10.495526v1

7. Merchlinsky M, Albright A, Olson V, Schiltz H, Merkeley T, Hughes C, et al. The development and approval of tecoviromat (TPOXX®), the first antiviral against smallpox. Antiviral Res. 2019 Aug;168:168–74.

8. Hatcher EL, Wang C, Lefkowitz EJ. Genome variability and gene content in chordopoxviruses: dependence on microsatellites. Viruses. 2015 Apr 22;7(4):2126–46.

